# Genotypes and associations with symptoms in primary ciliary dyskinesia

**DOI:** 10.1101/2023.11.28.23299010

**Authors:** Eva SL Pedersen, Myrofora Goutaki, Leonie D Schreck, Bernhard Rindlisbacher, Lucy Dixon, COVID-PCD patient advisory group, Jane S Lucas, Claudia E Kuehni

**Affiliations:** Institute of Social and Preventive Medicine, University of Bern, Bern, Switzerland; Division of Paediatric Respiratory Medicine and Allergology, Department of Paediatrics, Inselspital, University Hospital, University of Bern, Switzerland; Graduate School for Health Sciences, University of Bern, Switzerland; Selbsthilfegruppe Primäre Ciliäre Dyskinesie, Steffisburg, Switzerland; PCD Support UK, Buckingham, United Kingdom; Primary Ciliary Dyskinesia Centre, NIHR Biomedical Research Centre, University Hospital Southampton NHS Foundation Trust, Southampton, United Kingdom; University of Southampton Faculty of Medicine, School of Clinical and Experimental Medicine, Southampton, United Kingdom; Associazione Italiana Discinesia Ciliare Primaria Sindrome di Kartagener Onlus Italy; Association ADCP, France; PCD Support UK; Asociación Española de Pacientes con Discinesia Ciliar Primaria, Spain; Selbsthilfegruppe Primäre Ciliäre Dyskinesie, Switzerland; PCD Australia Primary Ciliary Dyskinesia, Australia; PCD Foundation, USA; Verein Kartagener Syndrom und Primäre Ciliäre Dyskinesie, Germany

## Abstract

**Background:** Knowledge about genotype-phenotype associations is crucial for understanding the clinical variability of primary ciliary dyskinesia (PCD). We studied how feasible it is to collect information about causative genes directly from people with PCD through questionnaires, and investigated associations between clinical characteristics, symptoms, and genotype.

**Methods:** We used data from the anonymous international participatory cohort COVID-PCD, set up in 2020 to follow people with PCD during the COVID-19 pandemic. A baseline questionnaire asked genetic test results, clinical characteristics, and current symptoms. We grouped reported causative genes into categories based on associated defects and studied differences between groups.

**Results:** Among the 759 COVID-PCD study participants, 444 (58%) reported genetic testing, and of these, 289 (65%) reported that a gene was identified. We included 206 who knew and reported a causative gene. The most common genes were *DNAH5* (n=71; 34%), *DNAH11* (n=27; 13%), *CCDC40* (n=21; 10%), *DNAI1* (n=18; 9%), *CCDC39* (n=13; 6%), and RSPH1 (n=8; 4%). The dynein structure (DS) group was the largest (n=127) followed by the nexin-dynein regulatory complex (ND-RC) group (n=38), dynein assembly (DA) group (n=21), and radial spoke and central complex (RS-CC, n=20) Current age and sex were similar across groups; but median age at diagnosis was markedly higher in the RS-CC group (11 years) compared to 4–7 years in the other groups (p=0.035). Laterality defects were reported by one person (5%) in RS-CC group, compared with 37%-60% in other groups (p=0.001). Overall, symptoms were frequently reported by participants in all 4 groups with little difference between groups.

**Conclusion:** Our results confirmed known differences in laterality defects and congenital heart disease between genotypes and showed frequent upper and lower respiratory symptoms in all groups regardless of reported gene.

## Manuscript

Knowledge about genotype-phenotype associations is crucial for understanding the clinical variability of primary ciliary dyskinesia (PCD) [1]. More than 50 known disease-causing genes lead to different ciliary structural defects resulting in differing phenotypical characteristics [2]. For example, laterality defects are not associated with mutations in genes related with the central pair apparatus, nexin link, or radial spoke complex (e.g. *HYDIN* and *RSPH9*), while they are seen in approximately 50% of individuals with mutations in most other genes [3, 4]. Similarly, congenital heart disease is less frequent in patients with these genes as congenital heart disease is associated with laterality defects [5, 6]. Several studies investigated associations of lung function with genotype and reported steeper decline in forced expiratory volume in one second (FEV_1_) for people with mutations affecting the nexin-dynein regulatory complex (e.g. *CCDC39* and *CCDC40*) compared to mutations in other genes [7-9].

Few studies investigated differences in symptoms between genotypes with inconsistent results. Data from 118 children from North America showed little difference in prevalence of chronic cough, chronic nasal congestion, and neonatal respiratory distress between groups of different genotypes [10]. Data from 199 patients from the United Kingdom (UK) in contrast showed lower prevalence of neonatal respiratory distress among patients with causative genes affecting the dynein structure compared to other genotypes [9].

Large studies including both detailed patient-reported symptom data and genetic data from medical records can be difficult to design. We therefore studied first how feasible it is to collect information about causative genes directly from people with PCD through questionnaires, and second, investigated associations between clinical characteristics, symptoms, and genotype.

We used data from the anonymous international participatory cohort COVID-PCD—a study set up in 2020 to follow people with PCD during the COVID-19 pandemic and beyond (clinicaltrials.gov: NCT04602481) [11, 12]. The study is online and includes people of any age worldwide. Study participants registered through the study website (www.covid19pcd.ispm.ch) and provided online consent (ethical approval from the Cantonal Ethics Committee of Bern, Switzerland, Study ID: 2020-00830). The baseline questionnaire asked about diagnostic tests including genetic analysis, whether participants knew results, and if yes, which causative gene was found. We also asked about age at diagnosis, laterality defects, and congenital heart disease, and detailed questions about symptoms during the past 3 months [13].

We grouped reported genes into categories based on associated defects: 1) dynein structure (DS), including *DNAH5, DNAH11, DNAI1, DNAI2, ODAD2, DNAH9, ODAD4, ODAD1 (CCDC114)*; 2) dynein assembly (DA), including *CCDC103, DNAAF4, LRRC6 (DNAAF11), DNAAF3, SPAG1 (DNAAF13), ZYMND10 (DNAAF7), DNAAF5, CFAP300 (DNAAF17)*; 3) microtubular stabilization/nexin-dynein regulatory complex (ND-RC), including *CCDC39, CCDC40, CCDC65 (DRC2), DRC1*; 4) radial spoke and central complex (RS-CC), including *RSPH4A, RSPH1, HYDIN, RSPH9, RSPH3*; 5) and other function (*RPGR, CCNO, MCIDAS*) [9]. We also compared differences for single causative genes reported by more than 5 people.

Our study included 759 people with suspected or confirmed PCD with median age of 28 years (interquartile range 13–44, range 1–86), 452 (60%) were females. Participants came from 49 countries; most from the United States (n=132), England (n=128), Germany (n=111), Italy (n=52), and Switzerland (n=50).

In total, 444 (58%) reported genetic testing; 52 (11%) reported no mutation found; 103 (24%) either did not know or awaited results; and 289 (65%) reported a gene was identified. Among these, 229 (79%) participants knew and reported the causative gene. We excluded 23 participants with heterozygous mutations in different genes leaving 206 in our analysis. The most common genes were *DNAH5* (n=71; 34%), *DNAH11* (n=27; 13%), *CCDC40* (n=21; 10%), *DNAI1* (n=18; 9%), *CCDC39* (n=13; 6%), and RSPH1 (n=8; 4%).

The dynein structure (DS) group was the largest (n=127) followed by the nexin-dynein regulatory complex (ND-RC) group (n=38), dynein assembly (DA) group (n=21), and radial spoke and central complex (RS-CC, n=20) (Table 1). Current age and sex were similar across groups; but median age at diagnosis was markedly higher in the RS-CC group (11 years) compared to 4–7 years in the other groups (p=0.035). Laterality defects were reported by one person (5%) in RS-CC group, compared with 37%-60% in other groups (p=0.001). Overall, symptoms were frequently reported by participants in all 4 groups with little difference between groups (Table 1). Prevalence of cough (reported either daily, often, or sometimes during the past 3 months) ranged from 84 to 92% (p=0.474); ear pain between 33 and 43% (p=0.772); and shortness of breath between 45 and 62% (p=0.589) across genotype groups. The biggest difference was seen for chronic nose symptoms which was reported by 62% in the DA group and 86% in the DS group (p=0.064).

**Table 1:**
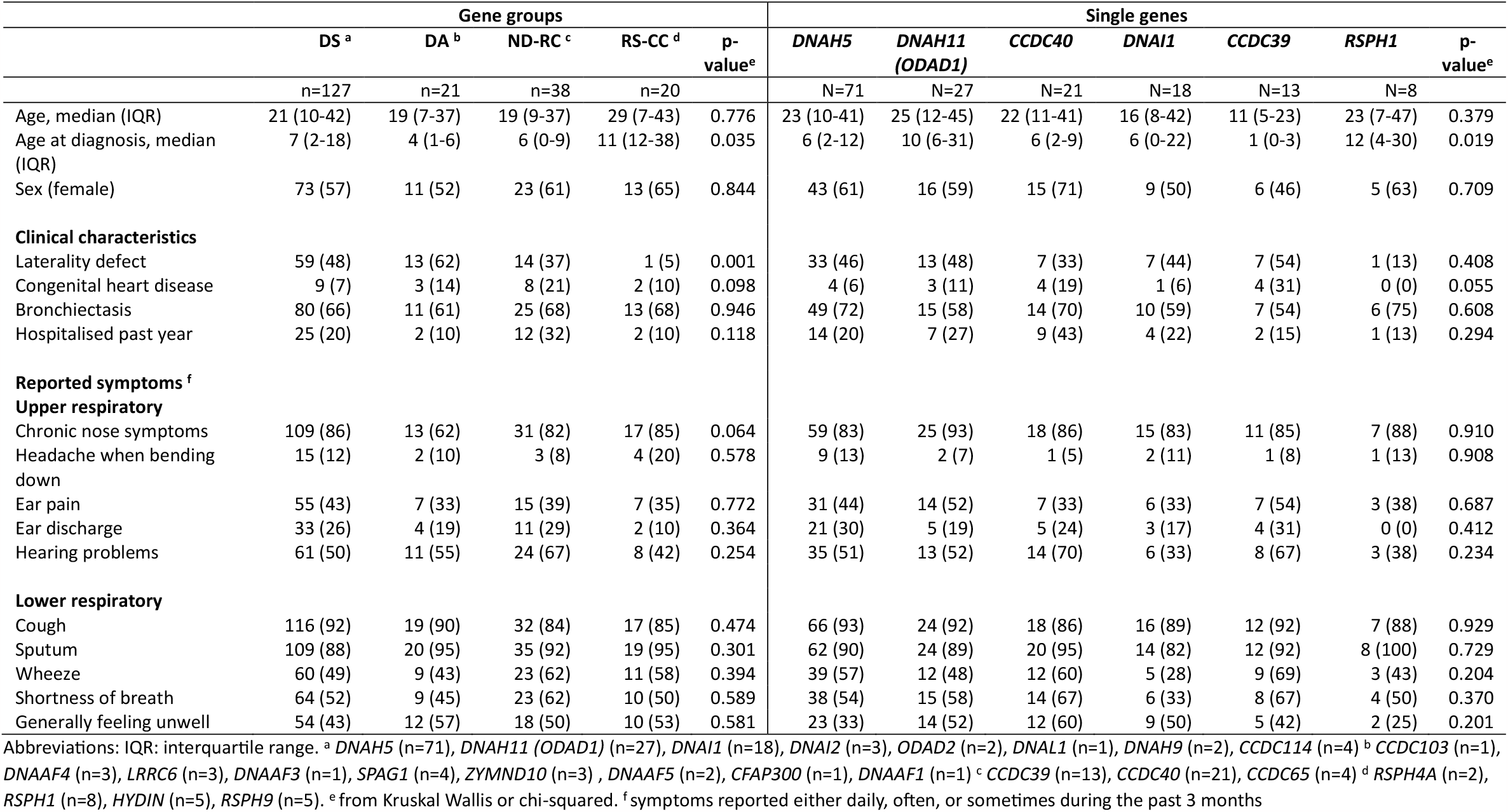
Clinical characteristics and symptoms by genotype groups including dynein structure (DS), dynein assembly (DA), nexin-dynein regulatory complex (ND-RC), and radial spoke and central complex (RS-CC) (n=206), and by single genes including *DNAH5, DNAH11 (ODAD1), CCDC40, DNAI1, CCDC39*, and *RSPH1* (n=158).

|  | Gene groups |  |  |  |  | Single genes |  |  |  |  |  |  |
| --- | --- | --- | --- | --- | --- | --- | --- | --- | --- | --- | --- | --- |
|  | DS <sup>a</sup> | DA <sup>b</sup> | ND-RC <sup>c</sup> | RS-CC <sup>d</sup> | p-value<sup>e</sup> | *DNAH5* | *DNAH11*<br>(*ODAD1*) | *CCDC40* | *DNAI1* | *CCDC39* | *RSPH1* | p-value<sup>e</sup> |
|  | n=127 | n=21 | n=38 | n=20 |  | N=71 | N=27 | N=21 | N=18 | N=13 | N=8 |  |
| Age, median (IQR) | 21 (10-42) | 19 (7-37) | 19 (9-37) | 29 (7-43) | 0.776 | 23 (10-41) | 25 (12-45) | 22 (11-41) | 16 (8-42) | 11 (5-23) | 23 (7-47) | 0.379 |
| Age at diagnosis, median (IQR) | 7 (2-18) | 4 (1-6) | 6 (0-9) | 11 (12-38) | 0.035 | 6 (2-12) | 10 (6-31) | 6 (2-9) | 6 (0-22) | 1 (0-3) | 12 (4-30) | 0.019 |
| Sex (female) | 73 (57) | 11 (52) | 23 (61) | 13 (65) | 0.844 | 43 (61) | 16 (59) | 15 (71) | 9 (50) | 6 (46) | 5 (63) | 0.709 |
| **Clinical characteristics** |  |  |  |  |  |  |  |  |  |  |  |  |
| Laterality defect | 59 (48) | 13 (62) | 14 (37) | 1 (5) | 0.001 | 33 (46) | 13 (48) | 7 (33) | 7 (44) | 7 (54) | 1 (13) | 0.408 |
| Congenital heart disease | 9 (7) | 3 (14) | 8 (21) | 2 (10) | 0.098 | 4 (6) | 3 (11) | 4 (19) | 1 (6) | 4 (31) | 0 (0) | 0.055 |
| Bronchiectasis | 80 (66) | 11 (61) | 25 (68) | 13 (68) | 0.946 | 49 (72) | 15 (58) | 14 (70) | 10 (59) | 7 (54) | 6 (75) | 0.608 |
| Hospitalised past year | 25 (20) | 2 (10) | 12 (32) | 2 (10) | 0.118 | 14 (20) | 7 (27) | 9 (43) | 4 (22) | 2 (15) | 1 (13) | 0.294 |
| **Reported symptoms <sup>f</sup>** |  |  |  |  |  |  |  |  |  |  |  |  |
| **Upper respiratory** |  |  |  |  |  |  |  |  |  |  |  |  |
| Chronic nose symptoms | 109 (86) | 13 (62) | 31 (82) | 17 (85) | 0.064 | 59 (83) | 25 (93) | 18 (86) | 15 (83) | 11 (85) | 7 (88) | 0.910 |
| Headache when bending down | 15 (12) | 2 (10) | 3 (8) | 4 (20) | 0.578 | 9 (13) | 2 (7) | 1 (5) | 2 (11) | 1 (8) | 1 (13) | 0.908 |
| Ear pain | 55 (43) | 7 (33) | 15 (39) | 7 (35) | 0.772 | 31 (44) | 14 (52) | 7 (33) | 6 (33) | 7 (54) | 3 (38) | 0.687 |
| Ear discharge | 33 (26) | 4 (19) | 11 (29) | 2 (10) | 0.364 | 21 (30) | 5 (19) | 5 (24) | 3 (17) | 4 (31) | 0 (0) | 0.412 |
| Hearing problems | 61 (50) | 11 (55) | 24 (67) | 8 (42) | 0.254 | 35 (51) | 13 (52) | 14 (70) | 6 (33) | 8 (67) | 3 (38) | 0.234 |
| **Lower respiratory** |  |  |  |  |  |  |  |  |  |  |  |  |
| Cough | 116 (92) | 19 (90) | 32 (84) | 17 (85) | 0.474 | 66 (93) | 24 (92) | 18 (86) | 16 (89) | 12 (92) | 7 (88) | 0.929 |
| Sputum | 109 (88) | 20 (95) | 35 (92) | 19 (95) | 0.301 | 62 (90) | 24 (89) | 20 (95) | 14 (82) | 12 (92) | 8 (100) | 0.729 |
| Wheeze | 60 (49) | 9 (43) | 23 (62) | 11 (58) | 0.394 | 39 (57) | 12 (48) | 12 (60) | 5 (28) | 9 (69) | 3 (43) | 0.204 |
| Shortness of breath | 64 (52) | 9 (45) | 23 (62) | 10 (50) | 0.589 | 38 (54) | 15 (58) | 14 (67) | 6 (33) | 8 (67) | 4 (50) | 0.370 |
| Generally feeling unwell | 54 (43) | 12 (57) | 18 (50) | 10 (53) | 0.581 | 23 (33) | 14 (52) | 12 (60) | 9 (50) | 5 (42) | 2 (25) | 0.201 |

Abbreviations: IQR: interquartile range. <sup>a</sup> *DNAH5* (n=71), *DNAH11* (*ODAD1*) (n=27), *DNAI1* (n=18), *DNAI2* (n=3), *ODAD2* (n=2), *DNAL1* (n=1), *DNAH9* (n=2), *CCDC114* (n=4) <sup>b</sup> *CCDC103* (n=1), *DNAAF4* (n=3), *LRRC6* (n=3), *DNAAF3* (n=1), *SPAG1* (n=4), *ZYMND10* (n=3), *DNAAF5* (n=2), *CFAP300* (n=1), *DNAAF1* (n=1) <sup>c</sup> *CCDC39* (n=13), *CCDC40* (n=21), *CCDC65* (n=4) <sup>d</sup> *RSPH4A* (n=2), *RSPH1* (n=8), *HYDIN* (n=5), *RSPH9* (n=5). <sup>e</sup> from Kruskal Wallis or chi-squared. <sup>f</sup> symptoms reported either daily, often, or sometimes during the past 3 months

Our analysis comparing single genes similarly showed difference in age at diagnosis with lowest age in participants with *CCDC39* (1 year) and highest age of diagnosis in participants with *RSPH1* (12 years) (p-value = 0.019). Laterality defect was reported in one person with *RSPH1* (13%) and between 33% and 54% in all other groups (p-value = 0.408). Congenital heart defect ranged from 0% in *RSPH1* to 31% in *CCDC39* (p=0.055). Again, symptoms were frequent across single genes.

This study using data contributed directly by people with PCD demonstrates that it is feasible to collect genetic information at the gene level by questionnaire. Among participants where a genetic cause was found, four out of five participants were able to report their causative gene. In line with predicted causative gene prevalence worldwide, we found that *DNAH5, DNAH11, and CCDC40* were most frequent [14]. Our findings reinforce previously shown genotypic differences. Laterality defect was not associated with RS-CC group in line with other studies [4, 15]. This may explain that age at diagnosis was highest for the RS-CC group, as people without laterality defects are often diagnosed later [16]. We found a higher prevalence of congenital heart disease for *CCDC39* and *CCDC40* than previously reported for PCD [4, 5, 9], there is however no mechanism to explain this finding. We found that study participants reported frequent symptoms independent of genotype or single gene suggesting that from a patient perspective, no phenotype without frequent symptoms exists, although some genotypes such as *DNAH11*, are considered milder and associated with preserved lung function [9].

The major strengths of our study include the sample size with 206 people and the combination of standardised patient-reported symptoms and genetics data, which allowed investigating novel genotype-phenotype associations. Our study is limited by our use of self-reported genetic data, which cannot be validated through linkage with clinical records from our anonymous study design. Without detailed clinical data on the mutation level, which patients usually do not have, we cannot be certain that all reported variants were pathogenic and that no single heterozygous variants or variants of unknown significance were reported as pathogenic. However, our data led to similar associations as previous studies which had used genetic information from clinics, suggesting patient-reported genetic information as reliable for epidemiological studies. Our results confirmed known differences in laterality defects and congenital heart disease between genotypes and showed frequent upper and lower respiratory symptoms in all groups regardless of reported gene.

## Data Availability

All data produced in the present study are available upon reasonable request to the authors

## Acknowledgements

We thank all participants and their families, and we thank the PCD support groups and physicians who advertised the study. We thank our collaborators who helped set up the COVID-PCD study: Cristina Ardura, Yin Ting Lam, Christina Mallet, Helena Koppe, Lara Pissini Goncalves from the University of Bern, and Amanda Harris from the University Hospital Southampton. We thank Kristin Bivens from the University of Bern for editing this research letter.

